# Causes of Death among Cancer Patients: Emerging Trends in the 21st Century

**DOI:** 10.1101/2024.10.18.24315724

**Authors:** Junpu Wang, Jie Liu, Ziyu Liu, Zongjiang Zhou, Diabate Ousmane, Liu Liu, Li Peng, Xiaoqing Yuan

## Abstract

Most studies examining the causes of cancer-related deaths have primarily focused on specific cancer types, often neglecting the evolving spectrum of death causes among cancer patients in the 21st century. This study, utilizing data from the National Cancer Institute’s Surveillance, Epidemiology, and End Results (SEER) Program, analyzed the causes of death in patients diagnosed with 36 types of cancer between 2000 and 2021. By categorizing these causes into deaths from index cancers, non-index cancers, and non-cancer causes, this study provides a comprehensive analysis of cause-of-death patterns and emerging trends, stratified by year of death, age at diagnosis, and survival duration. The findings reveal that while relative mortality rates from index cancers remain elevated for brain, pancreatic, and gallbladder cancers, significant declines were observed for lung, liver, nasopharyngeal, and esophageal cancers, as well as multiple myeloma cancers, reflecting advancements in cancer treatment. Besides, relative mortality rates from non-index cancers surpassed those from index cancers in oral cavity, oropharyngeal, vaginal, and small intestine cancers, indicating a potential benefit from enhanced surveillance and early detection of non-index cancers in these patient populations. Importantly, non-cancer-related causes of death, such as heart disease, chronic liver disease and cirrhosis, also emerged as prominent contributors to mortality among cancer patients. The results of this study offer critical and current data to inform public health policy, optimize healthcare resource allocation, and facilitate international collaboration in cancer research and control. Meanwhile, this study is of great reference value for developing countries to formulate medium– and long-term public health policies.

## Introduction

Cancer remains one of the most significant threats to human health worldwide, imposing severe physical and psychological burdens on patients and their families, while also challenging healthcare systems and public health policies^1^. In 2022, nearly 20 million new cancer cases and 9.7 million cancer-related deaths were reported globally^1^. Despite advances in medical technology and the emergence of new cancer treatments^2–6^, survival rates among cancer patients have significantly improved^7^. However, cancer continues to be one of the leading causes of death in many countries, with millions of individuals succumbing to the disease each year^8^. Understanding the causes of death among cancer patients is critical not only for improving their survival rates and quality of life but also for guiding the development of prevention and treatment strategies for cancer-related mortality and advancing basic and translational research into these causes.

The causes of death among cancer patients have been partially explored in previous epidemiological studies. Using U.S. cancer data from 1973 to 2012, Zaorsky et al. classified the causes of death among cancer patients into three categories: index cancer, non-index cancer, and non-cancer causes^9^. This study comprehensively analyzed the risk of death from these three categories across 28 cancer types, comparing the risk of death from 13 non-cancer causes between cancer patients and the general population. These findings provided valuable insights into trends in causes of death among cancer patients over time^9^. Additionally, some studies have revealed generational differences in the causes of death in cancer patients. For instance, early research identified metastasis as the leading cause of death in breast cancer patients^10^. However, with advances in medical technology and improved treatments, researchers are increasingly recognizing that other factors also significantly influence the causes of death among cancer patients. For example, Ramin et al. reported that cardiovascular disease accounted for a considerable proportion of deaths among breast cancer patients^11^. Although these studies have enhanced our understanding of cancer mortality, several limitations remain. First, most studies have concentrated on specific cancer types, leading to an incomplete understanding of the causes of death across all cancer patients. Second, there is a scarcity of research examining shifts in the spectrum of death causes among cancer patients during the first two decades of the 21st century.

In this context, the objective of this study is to provide a detailed analysis of the causes of death among cancer patients, focusing on the first two decades of the 21st century. Using data from the SEER database, the study will categorize death causes by year of death, age at diagnosis, and survival time after diagnosis. The goal is to supply healthcare systems with the most current data and trend analyses on cancer mortality. The results of this study will offer valuable insights for formulating public health policies, optimizing medical resource allocation, fostering international collaboration and knowledge exchange in cancer research, prevention, and control, and enhancing public awareness of cancer-related mortality.

## Materials and Methods

### Study Design and Data Source

Data for this observational study were obtained from the National Cancer Institute’s SEER program, a population-based cancer registry covering approximately 47.9% of the U.S. population. The SEER database regularly collects and publishes detailed information on cancer incidence, survival rates, treatment modalities, and causes of death^12^. SEER*Stat software (version 8.4.3) was used to extract data on cancer patients from 2000 to 2021, specifically from the SEER 17 registry (2023 submission), which includes approximately 26.5% of the U.S. population^13–15^. Detailed session information for extracting cancer data is available in the supplementary material S1. As SEER data are anonymized and classified as non-human subject research, this study did not require institutional review board approval. The study adhered to the Strengthening the Reporting of Observational Studies in Epidemiology (STROBE) guidelines^16^.

### Study Population

The study included all patients diagnosed with malignant tumors between January 1, 2000, and December 31, 2021. To ensure the focus was on patients’ first cancer diagnoses, only cases with “one primary only” and “1st of 2 or more primaries” tumor sequence numbers were included. Patients whose cancer diagnoses were made solely through autopsy or death certificate were excluded. For comparison with the general population, mortality data for the general U.S. population from 2000 to 2021 were obtained from the National Center for Health Statistics through the SEER program.

### Definition of Variables

Cancer classifications were defined according to SEER standards, based on the International Classification of Diseases for Oncology, Third Edition (ICD-O-3). Cause-of-death information was sourced from death certificates and defined by SEER using the International Classification of Diseases, 10th Edition (ICD-10). Causes of death were categorized into three primary groups: index cancer deaths, non-index cancer deaths, and non-cancer deaths. Index cancer deaths refer to deaths resulting directly from the initially diagnosed primary cancer. Non-index cancer deaths refer to deaths caused by another independent cancer unrelated to the initial diagnosis. Non-cancer deaths encompass deaths from non-cancer-related medical causes, grouped into 25 categories (supplementary Table S1). In some instances, primary cancer deaths may have been miscoded; to address this, deaths from certain sites were reclassified as index cancer deaths if attributed to the local or regional spread of the primary cancer. For example, if a patient initially diagnosed with pancreatic cancer was coded as having died from “liver cancer”, the cause of death was recategorized as pancreatic cancer (supplementary Table S2). Post-diagnostic follow-up time was defined as the interval from cancer diagnosis to death from any cause, the date of the last follow-up, or the end of the study on December 31, 2021.

### Statistical Analysis

Relative mortality rates for causes of death were calculated as the ratio of total deaths during the follow-up period to the number of deaths attributed to specific causes. For non-cancer causes, standardized mortality ratios (SMRs) were calculated^17^. The SMR is based on the assumption that the general population is cancer-free, comparing mortality rates among cancer patients to those in the general population to assess whether cancer patients face an elevated risk of death from specific non-cancer causes. SMRs were computed as the ratio of observed to expected deaths, where observed deaths reflect the actual number of deaths among cancer patients from a specific cause, and expected deaths represent the number of deaths from the same cause in a demographically matched general population (e.g., age, sex, race, and calendar year). The Poisson distribution method was used to calculate 95% confidence intervals for SMRs, and all statistical tests were two-sided with P-values <0.05 considered statistically significant. Analyses were performed using SEER*Stat (version 8.4.3) and Microsoft Excel 2021 (version 2407, build 16.0.17830.20056, 64-bit)^14^.

## Results

### Baseline Characteristics

Between 2000 and 2021, a total of 6,903,506 patients were diagnosed with cancer, of whom 3,364,903 (48.7%) had died by December 31, 2021. The median age at cancer diagnosis was 65.1 years. Age at diagnosis varied across tumor types, with testicular cancer being diagnosed at a younger median age (33.3 years) and bladder cancer at an older median age (71.3 years). The overall gender distribution among cancer patients was similar (males: 51.1%); however, in addition to cancers that are exclusively gender-specific, there were substantial gender differences for certain cancers. Nearly all breast cancer patients were female (99.3%), whereas bladder cancer predominantly affected males (75.6%). Additionally, the majority of patients were white. Detailed baseline characteristics are presented in Table 1.

**Table 1.** Demographic characteristics and number of deaths among patients diagnosed with cancer between 2000 and 2021.

| Cancer site | Age at | Sex |  |  |  | Race |  |  |  |  |  |  |  |  |  | Vital status |  |  |  |
| --- | --- | --- | --- | --- | --- | --- | --- | --- | --- | --- | --- | --- | --- | --- | --- | --- | --- | --- | --- |
|  | diagnosis | Men |  | Women |  | White |  | Black |  | AI/AN |  | API |  | Unknown |  | Alive |  | Death |  |
|  | Median | cases | %<sup>a</sup> | cases | %<sup>a</sup> | cases | %<sup>a</sup> | cases | %<sup>a</sup> | cases | %<sup>a</sup> | cases | %<sup>a</sup> | cases | %<sup>a</sup> | cases | %<sup>a</sup> | cases | %<sup>a</sup> |
| All sites | 65.1 | 3530407 | 51.1 | 3373099 | 48.9 | 5557818 | 80.5 | 713921 | 10.3 | 43763 | 0.6 | 495284 | 7.2 | 92720 | 1.3 | 3538603 | 51.3 | 3364903 | 48.7 |
| Lip, oral cavity | 67.2 | 59597 | 67.6 | 28619 | 32.4 | 75046 | 85.1 | 6141 | 7.0 | 511 | 0.6 | 5309 | 6.0 | 1209 | 1.4 | 43765 | 49.6 | 44451 | 50.4 |
| Salivary gland | 63.4 | 10457 | 57.7 | 7674 | 42.3 | 14450 | 79.7 | 1668 | 9.2 | 122 | 0.7 | 1652 | 9.1 | 239 | 1.3 | 10383 | 57.3 | 7748 | 42.7 |
| Nasopharynx | 55.5 | 7285 | 70.5 | 3051 | 29.5 | 4778 | 46.2 | 1197 | 11.6 | 165 | 1.6 | 4078 | 39.5 | 118 | 1.1 | 5182 | 50.1 | 5154 | 49.9 |
| Tonsil | 58.7 | 26472 | 83.2 | 5334 | 16.8 | 27695 | 87.1 | 2638 | 8.3 | 229 | 0.7 | 1002 | 3.2 | 242 | 0.8 | 19623 | 61.7 | 12183 | 38.3 |
| Oropharynx | 61.9 | 5601 | 79.2 | 1468 | 20.8 | 5818 | 82.3 | 935 | 13.2 | 51 | 0.7 | 205 | 2.9 | 60 | 0.8 | 3093 | 43.8 | 3976 | 56.2 |
| Hypopharynx | 63.9 | 6695 | 80.7 | 1598 | 19.3 | 6365 | 76.8 | 1295 | 15.6 | 87 | 1.0 | 522 | 6.3 | 24 | 0.3 | 1791 | 21.6 | 6502 | 78.4 |
| Esophagus | 67.0 | 49589 | 78.4 | 13622 | 21.6 | 52837 | 83.6 | 6425 | 10.2 | 482 | 0.8 | 3241 | 5.1 | 226 | 0.4 | 10491 | 16.6 | 52720 | 83.4 |
| Stomach | 67.8 | 65300 | 60.0 | 43526 | 40.0 | 76176 | 70.0 | 14447 | 13.3 | 1103 | 1.0 | 16330 | 15.0 | 770 | 0.7 | 28616 | 26.3 | 80210 | 73.7 |
| Small intestine | 64.4 | 16061 | 52.5 | 14542 | 47.5 | 23523 | 76.9 | 4891 | 16.0 | 171 | 0.6 | 1714 | 5.6 | 304 | 1.0 | 16266 | 53.2 | 14337 | 46.8 |
| Colon | 68.6 | 217091 | 49.4 | 222770 | 50.6 | 345505 | 78.5 | 52882 | 12.0 | 3286 | 0.7 | 35049 | 8.0 | 3139 | 0.7 | 185826 | 42.2 | 254035 | 57.8 |
| Rectum | 62.1 | 83768 | 57.8 | 61052 | 42.2 | 111923 | 77.3 | 14774 | 10.2 | 1325 | 0.9 | 14608 | 10.1 | 2190 | 1.5 | 74762 | 51.6 | 70058 | 48.4 |
| Anus | 61.5 | 9776 | 36.9 | 16715 | 63.1 | 22654 | 85.5 | 2688 | 10.1 | 188 | 0.7 | 759 | 2.9 | 202 | 0.8 | 14553 | 54.9 | 11938 | 45.1 |
| Liver | 64.0 | 85563 | 74.5 | 29295 | 25.5 | 79947 | 69.6 | 13855 | 12.1 | 1673 | 1.5 | 18742 | 16.3 | 641 | 0.6 | 22822 | 19.9 | 92036 | 80.1 |
| Gallbladder | 70.8 | 5385 | 30.2 | 12444 | 69.8 | 13418 | 75.3 | 2109 | 11.8 | 267 | 1.5 | 1948 | 10.9 | 87 | 0.5 | 3157 | 17.7 | 14672 | 82.3 |
| Pancreas | 69.8 | 91191 | 50.4 | 89819 | 49.6 | 143818 | 79.5 | 20955 | 11.6 | 1210 | 0.7 | 14425 | 8.0 | 602 | 0.3 | 21209 | 11.7 | 159801 | 88.3 |
| Larynx | 64.4 | 37836 | 80.7 | 9061 | 19.3 | 37799 | 80.6 | 6898 | 14.7 | 266 | 0.6 | 1655 | 3.5 | 279 | 0.6 | 17910 | 38.2 | 28987 | 61.8 |
| Lung and<br>bronchus | 69.7 | 409532 | 52.7 | 366912 | 47.3 | 635673 | 81.9 | 82565 | 10.6 | 4409 | 0.6 | 51994 | 6.7 | 1803 | 0.2 | 121342 | 15.6 | 655102 | 84.4 |
| Bones and joints | 37.4 | 8288 | 56.4 | 6404 | 43.6 | 11826 | 80.5 | 1446 | 9.8 | 122 | 0.8 | 1130 | 7.7 | 168 | 1.1 | 9046 | 61.6 | 5646 | 38.4 |
| Melanoma of the | 60.5 | 172892 | 55.9 | 136355 | 44.1 | 288561 | 93.3 | 1334 | 0.4 | 798 | 0.3 | 2056 | 0.7 | 16498 | 5.3 | 231594 | 74.9 | 77653 | 25.1 |
| skin |  |  |  |  |  |  |  |  |  |  |  |  |  |  |  |  |  |  |  |
| Breast | 60.9 | 7348 | 0.7 | 1038205 | 99.3 | 829147 | 79.3 | 109029 | 10.4 | 6439 | 0.6 | 92929 | 8.9 | 8009 | 0.8 | 739704 | 70.7 | 305849 | 29.3 |
| Cervix uteri | 48.3 | 0 | 0.0 | 67376 | 100.0 | 50525 | 75.0 | 8908 | 13.2 | 663 | 1.0 | 6533 | 9.7 | 747 | 1.1 | 41756 | 62.0 | 25620 | 38.0 |
| Corpus uteri | 62.3 | 0 | 0.0 | 218759 | 100.0 | 174584 | 79.8 | 21200 | 9.7 | 1632 | 0.7 | 19393 | 8.9 | 1950 | 0.9 | 151033 | 69.0 | 67726 | 31.0 |
| Ovary | 62.4 | 0 | 0.0 | 99923 | 100.0 | 81731 | 81.8 | 8237 | 8.2 | 727 | 0.7 | 8713 | 8.7 | 515 | 0.5 | 38736 | 38.8 | 61187 | 61.2 |
| Vagina | 66.9 | 0 | 0.0 | 4453 | 100.0 | 3441 | 77.3 | 669 | 15.0 | 23 | 0.5 | 290 | 6.5 | 30 | 0.7 | 1753 | 39.4 | 2700 | 60.6 |
| Vulva | 67.0 | 0 | 0.0 | 17523 | 100.0 | 15095 | 86.1 | 1420 | 8.1 | 113 | 0.6 | 675 | 3.9 | 220 | 1.3 | 9010 | 51.4 | 8513 | 48.6 |
| Prostate | 67.1 | 1055784 | 100.0 | 0 | 0.0 | 813638 | 77.1 | 155148 | 14.7 | 4380 | 0.4 | 54426 | 5.2 | 28192 | 2.7 | 704753 | 66.8 | 351031 | 33.2 |
| Testis | 33.3 | 49002 | 100.0 | 0 | 0.0 | 43717 | 89.2 | 1438 | 2.9 | 545 | 1.1 | 2091 | 4.3 | 1211 | 2.5 | 44746 | 91.3 | 4256 | 8.7 |
| Penis | 66.3 | 5621 | 100.0 | 0 | 0.0 | 4697 | 83.6 | 504 | 9.0 | 54 | 1.0 | 284 | 5.1 | 82 | 1.5 | 2626 | 46.7 | 2995 | 53.3 |
| Urinary bladder | 71.3 | 200325 | 75.6 | 64606 | 24.4 | 234454 | 88.5 | 14171 | 5.3 | 1004 | 0.4 | 12074 | 4.6 | 3228 | 1.2 | 127324 | 48.1 | 137607 | 51.9 |
| Kidney, renal<br>pelvis | 63.1 | 140095 | 62.6 | 83587 | 37.4 | 182745 | 81.7 | 23991 | 10.7 | 2471 | 1.1 | 12643 | 5.7 | 1832 | 0.8 | 128735 | 57.6 | 94947 | 42.4 |
| Brain, other<br>nervous system | 55.5 | 54029 | 55.9 | 42592 | 44.1 | 83075 | 86.0 | 6511 | 6.7 | 591 | 0.6 | 5809 | 6.0 | 635 | 0.7 | 32068 | 33.2 | 64553 | 66.8 |
| Thyroid | 48.7 | 46235 | 23.5 | 150241 | 76.5 | 157112 | 80.0 | 12763 | 6.5 | 1402 | 0.7 | 22284 | 11.3 | 2915 | 1.5 | 176187 | 89.7 | 20289 | 10.3 |
| Hodgkin<br>lymphoma | 36.4 | 23647 | 54.7 | 19559 | 45.3 | 34952 | 80.9 | 5125 | 11.9 | 234 | 0.5 | 2358 | 5.5 | 537 | 1.2 | 33866 | 78.4 | 9340 | 21.6 |
| NK lymphoma | 65.3 | 155227 | 54.4 | 130119 | 45.6 | 236257 | 82.8 | 22308 | 7.8 | 1598 | 0.6 | 21275 | 7.5 | 3908 | 1.4 | 150436 | 52.7 | 134910 | 47.3 |
| Myeloma | 68.1 | 53886 | 54.7 | 44678 | 45.3 | 71882 | 72.9 | 19163 | 19.4 | 682 | 0.7 | 6067 | 6.2 | 770 | 0.8 | 37395 | 37.9 | 61169 | 62.1 |
| Leukemia | 63.9 | 113604 | 57.6 | 83766 | 42.4 | 165483 | 83.8 | 15681 | 7.9 | 1297 | 0.7 | 12028 | 6.1 | 2881 | 1.5 | 95812 | 48.5 | 101558 | 51.5 |

Note: vital status was followed until 31 December 2021.
Abbreviation: AI/AN, American Indian/Alaska Native. API, Asian or Pacific Islander. NK Lymphoma, Non-Hodgkin Lymphoma.
<sup>a</sup> Percentage of total cancer patients.

### Causes of Death: 2000-2021

#### Index-Cancer Deaths

From 2000 to 2021, the proportion of deaths attributed to index cancer steadily declined, decreasing from 78% in 2000 to 44.9% in 2021. In contrast, non-cancer-related deaths increased from 14.8% in 2000 to 43.4% in 2021 (Figure 1A). Lung cancer contributed the largest proportion of index cancer deaths (Figure 1B). Furthermore, in 2020, there was a sharp decline in the proportion of deaths caused by index cancer, accompanied by a corresponding increase in non-cancer causes of death (Figure 1A). This trend was observed across the majority of cancer types, including cancers of the salivary gland, nasopharynx, thyroid, breast, corpus uteri, vagina, vulva, colon, liver, lymphoma, multiple myeloma, leukemia, prostate, testis, bladder, kidney, and lung (Figure 2). Cancer types that showed the most significant declines in the proportion of index cancer deaths included thyroid cancer, kidney cancer, leukemia, testicular cancer, melanoma, non-Hodgkin lymphoma, bladder cancer, breast cancer, uterine corpus cancer, anal canal cancer, laryngeal cancer, nasopharyngeal cancer, vulva cancer, colon cancer, and Hodgkin lymphoma. By contrast, the proportion of deaths due to index cancer remained stable for pancreatic cancer, malignant tumors of the brain and central nervous system, oropharyngeal cancer, gallbladder cancer, and malignant tumors of the bone and joints (Figure 2, black lines).

**Figure 1.**
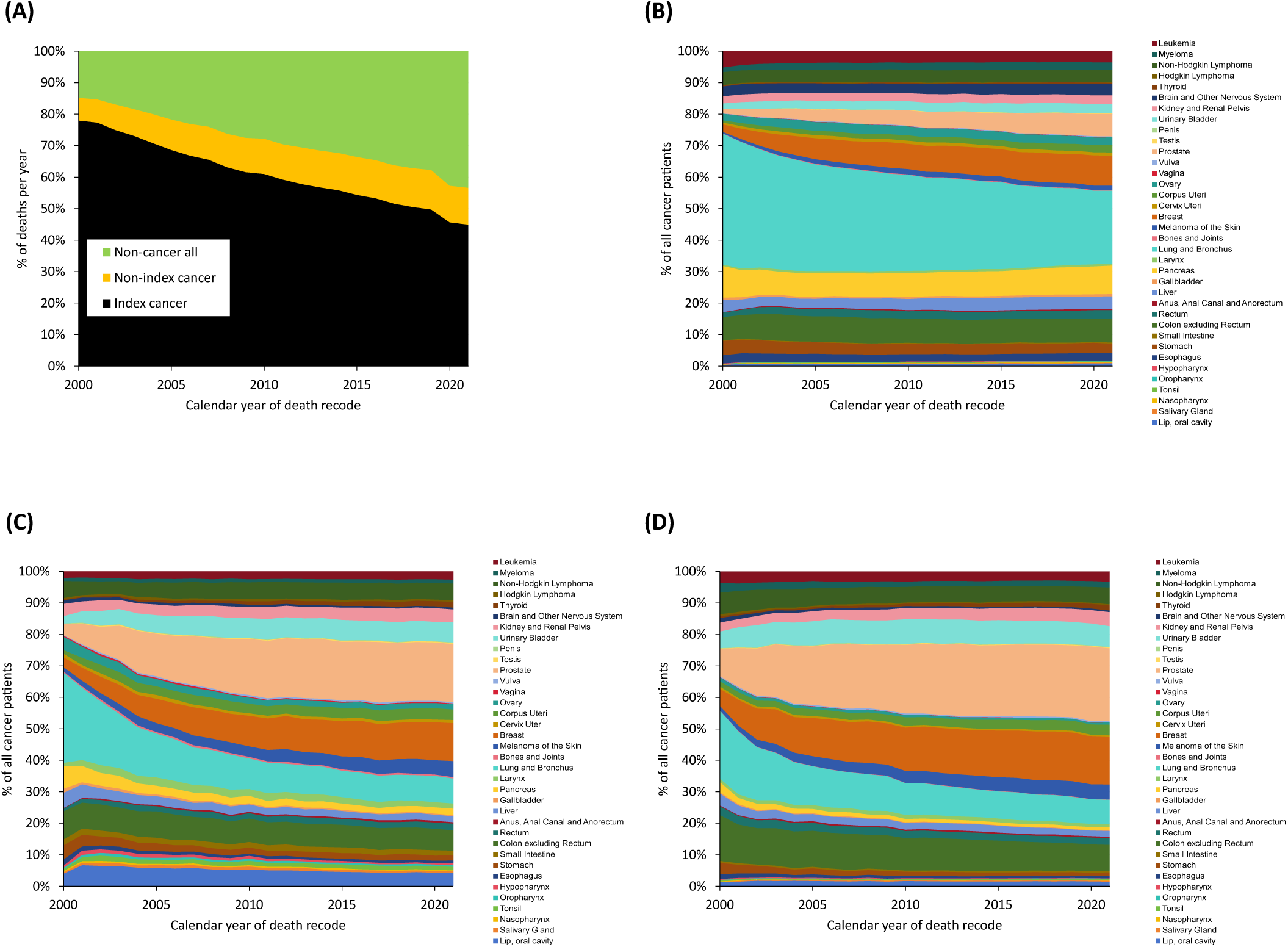
Overall trends in causes of death among cancer patients in the 21st century. (A) Proportional graph of cancer patients’ deaths due to index cancers (black), non-index cancers (yellow), and non-cancer causes (green) from 2000 to 2021. The proportion of index-cancer deaths has been continuously decreasing, while deaths due to non-cancer causes have increased. Proportional graph of deaths caused by (B) index cancers, (C) non-index cancers, and (D) non-cancer causes, stratified by various types of cancer.

**Figure 2.**
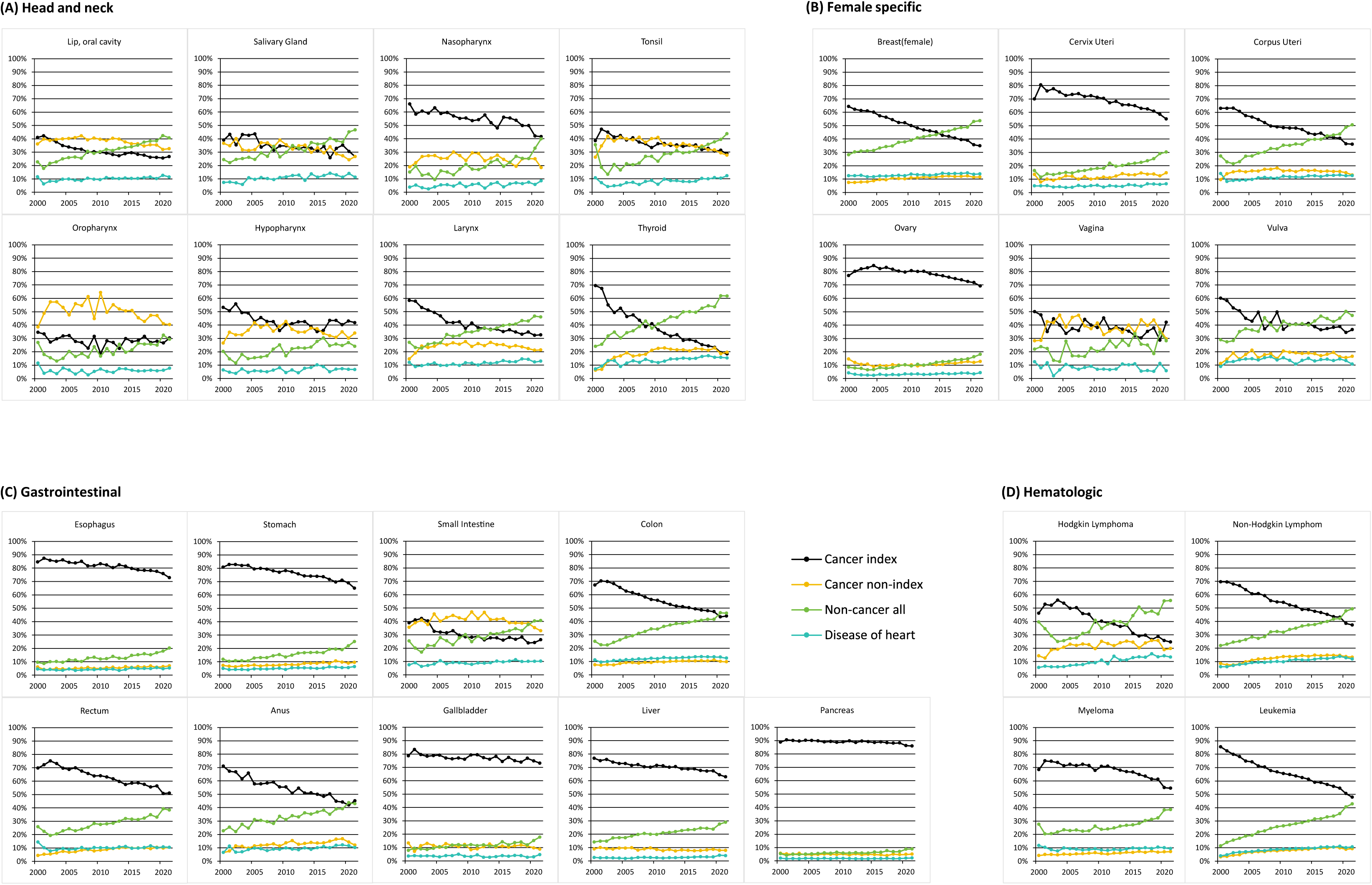
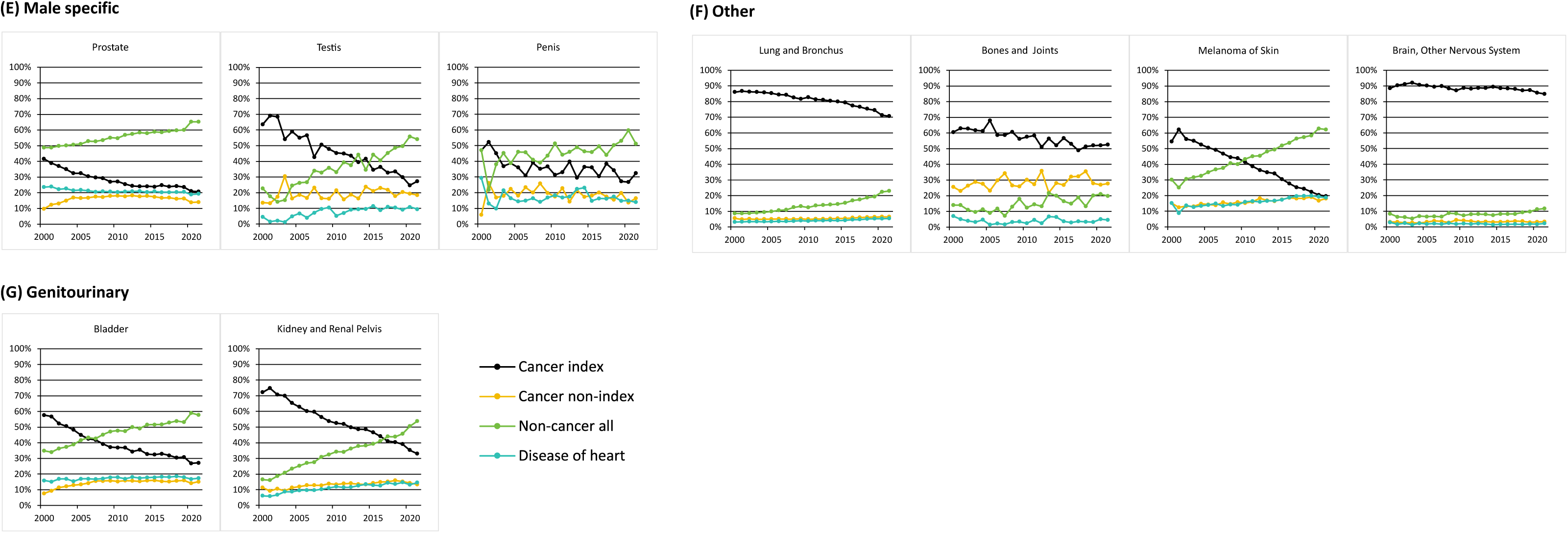
Proportion of cause of death versus year of death among cancer patients diagnosed between 2000 and 2021 for various cancers. Death was characterized as due to “index-cancer” (black lines), “non-index cancer” (yellow line), and “noncancer death” (green lines). Year of death refers of the year in which the death occurred.

#### Non-Index Cancer Deaths

Prostate and breast cancers accounted for a substantial proportion of non-index cancer deaths. Although the non-index cancer death rate for lung cancer was relatively low, the absolute number of non-index cancer deaths remained significant due to its high incidence and mortality (Figure 1C). The cancer types with the highest non-index cancer death rates included head and neck cancers, vaginal cancer, small intestine cancer, Hodgkin lymphoma, testicular cancer, penile cancer, and malignant tumors of the bone and joints. Thyroid cancer showed the largest increase in the non-index cancer death rate between 2000 and 2021, whereas other cancer types exhibited stable trends over time (Figure 2, yellow lines).

#### Non-Cancer Deaths

Among non-cancer causes of death, prostate and breast cancers were associated with the largest numbers of deaths, followed by lung and colon cancers (Figure 1D). Over time, for patients with cancers of the head and neck (including oral cavity, salivary glands, tonsils, larynx, and thyroid), breast, uterine corpus, vulva, small intestine, lymphoma, male genital tract, genitourinary tract, and melanoma, the proportion of deaths from non-cancer causes surpassed that from index cancers (Figure 2, green lines). This trend suggests that non-cancer causes are becoming the leading causes of death in these cancer populations. Heart disease was the most frequent cause of non-cancer-related deaths. A higher proportion of heart disease-related deaths was observed among patients with thyroid cancer, prostate cancer, penile cancer, bladder cancer, kidney cancer, and melanoma (Figure 2, blue lines). Between 2000 and 2021, 946,584 cancer patients died from non-cancer causes. The leading non-cancer causes of death were heart disease (305,634 deaths) and other causes (213,594 deaths) (Table 2). Non-cancer death rates varied by age group. Among patients under 45 years of age, the primary non-cancer causes of death included other causes, infectious and parasitic diseases, and HIV. Accidents and adverse effects were more prevalent among patients aged 20-39 years. For patients over 45 years of age, heart disease and other causes were the most common non-cancer causes of death (Figure 3A). Heart disease consistently accounted for the largest proportion of non-cancer deaths, regardless of follow-up duration (Figure 3B). A heat map comparing the 15 most common non-cancer causes of death showed that chronic liver disease and cirrhosis (SMR: 12.5 [95% CI: 12.3-12.7]), other infectious and parasitic diseases, HIV (SMR: 9.98 [95% CI: 9.84-10.12]), and suicide and self-inflicted injury (SMR: 8.71 [95% CI: 8.53-8.9]) had the highest SMRs (Supplementary material S1_SMR heatmap table).

**Figure 3.**
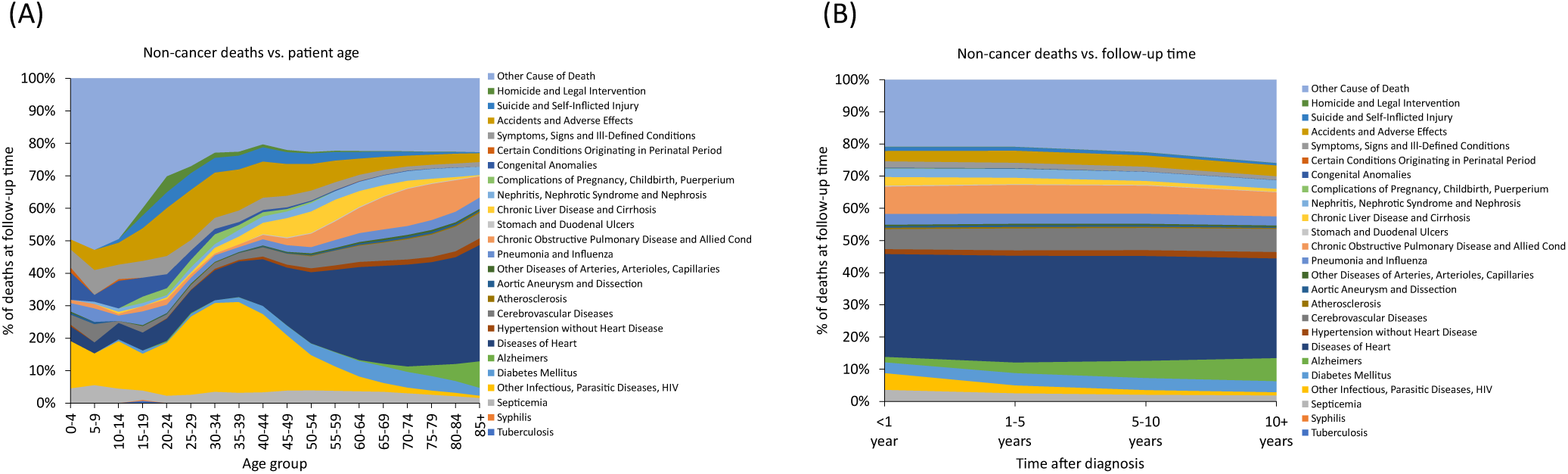
Overall trends of the proportion of non-cancer causes of death among cancer patients in the 21st century. Proportional graphs of death due to non-cancer causes among cancer patients diagnosed between 2000 and 2021 by (A) patient age and (B) time after diagnosis, stratified by various non-cancer causes.

**Table 2.**
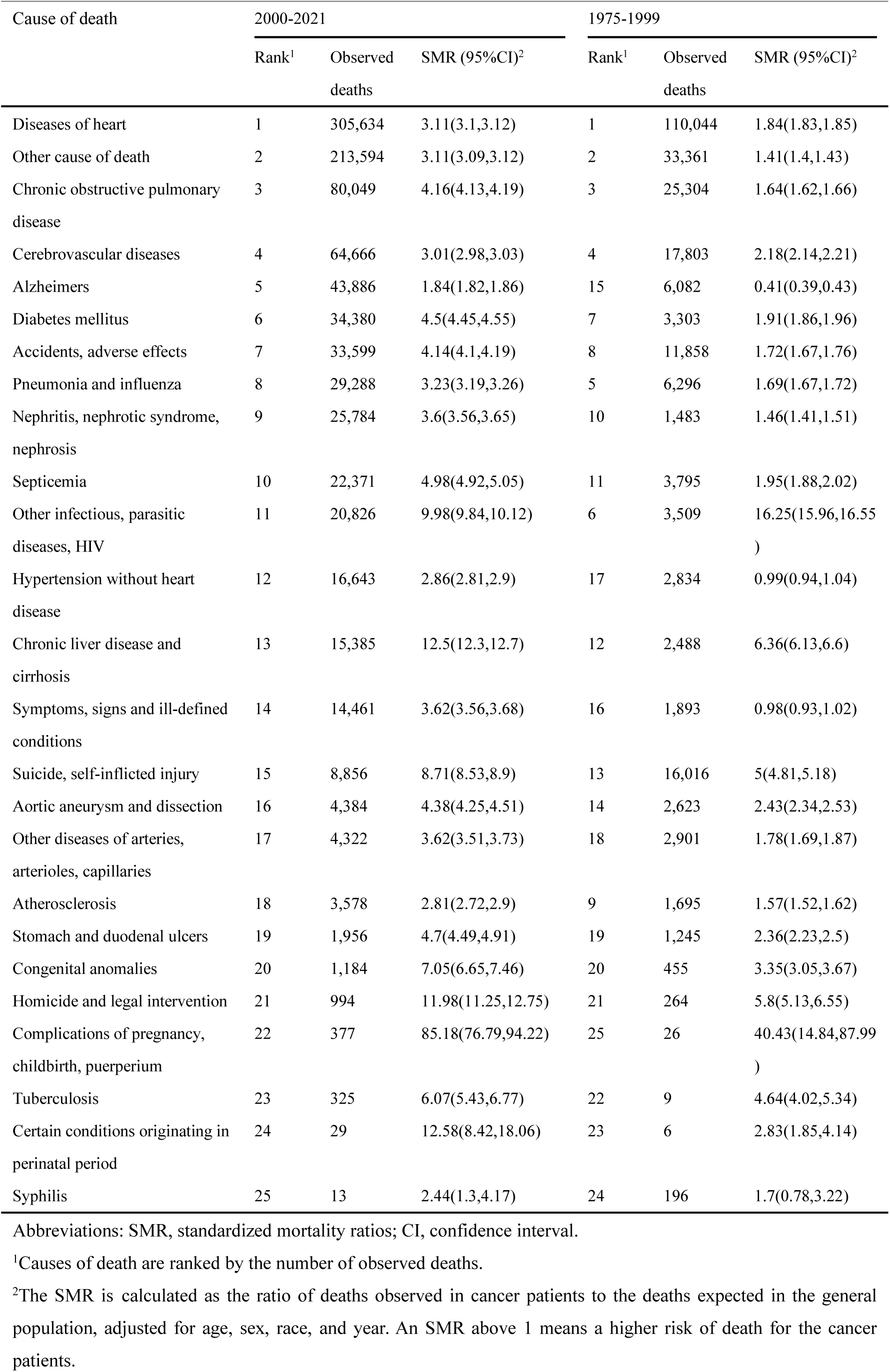
Standardized mortality ratios of non-cancer death among cancer patients.

| Cause of death | 2000-2021 |  |  | 1975-1999 |  |  |
| --- | --- | --- | --- | --- | --- | --- |
|  | Rank<sup>1</sup> | Observed deaths | SMR (95%CI)<sup>2</sup> | Rank<sup>1</sup> | Observed deaths | SMR (95%CI)<sup>2</sup> |
| Diseases of heart | 1 | 305,634 | 3.11(3.1,3.12) | 1 | 110,044 | 1.84(1.83,1.85) |
| Other cause of death | 2 | 213,594 | 3.11(3.09,3.12) | 2 | 33,361 | 1.41(1.4,1.43) |
| Chronic obstructive pulmonary disease | 3 | 80,049 | 4.16(4.13,4.19) | 3 | 25,304 | 1.64(1.62,1.66) |
| Cerebrovascular diseases | 4 | 64,666 | 3.01(2.98,3.03) | 4 | 17,803 | 2.18(2.14,2.21) |
| Alzheimers | 5 | 43,886 | 1.84(1.82,1.86) | 15 | 6,082 | 0.41(0.39,0.43) |
| Diabetes mellitus | 6 | 34,380 | 4.5(4.45,4.55) | 7 | 3,303 | 1.91(1.86,1.96) |
| Accidents, adverse effects | 7 | 33,599 | 4.14(4.1,4.19) | 8 | 11,858 | 1.72(1.67,1.76) |
| Pneumonia and influenza | 8 | 29,288 | 3.23(3.19,3.26) | 5 | 6,296 | 1.69(1.67,1.72) |
| Nephritis, nephrotic syndrome, nephrosis | 9 | 25,784 | 3.6(3.56,3.65) | 10 | 1,483 | 1.46(1.41,1.51) |
| Septicemia | 10 | 22,371 | 4.98(4.92,5.05) | 11 | 3,795 | 1.95(1.88,2.02) |
| Other infectious, parasitic diseases, HIV | 11 | 20,826 | 9.98(9.84,10.12) | 6 | 3,509 | 16.25(15.96,16.55) |
| Hypertension without heart disease | 12 | 16,643 | 2.86(2.81,2.9) | 17 | 2,834 | 0.99(0.94,1.04) |
| Chronic liver disease and cirrhosis | 13 | 15,385 | 12.5(12.3,12.7) | 12 | 2,488 | 6.36(6.13,6.6) |
| Symptoms, signs and ill-defined conditions | 14 | 14,461 | 3.62(3.56,3.68) | 16 | 1,893 | 0.98(0.93,1.02) |
| Suicide, self-inflicted injury | 15 | 8,856 | 8.71(8.53,8.9) | 13 | 16,016 | 5(4.81,5.18) |
| Aortic aneurysm and dissection | 16 | 4,384 | 4.38(4.25,4.51) | 14 | 2,623 | 2.43(2.34,2.53) |
| Other diseases of arteries, arterioles, capillaries | 17 | 4,322 | 3.62(3.51,3.73) | 18 | 2,901 | 1.78(1.69,1.87) |
| Atherosclerosis | 18 | 3,578 | 2.81(2.72,2.9) | 9 | 1,695 | 1.57(1.52,1.62) |
| Stomach and duodenal ulcers | 19 | 1,956 | 4.7(4.49,4.91) | 19 | 1,245 | 2.36(2.23,2.5) |
| Congenital anomalies | 20 | 1,184 | 7.05(6.65,7.46) | 20 | 455 | 3.35(3.05,3.67) |
| Homicide and legal intervention | 21 | 994 | 11.98(11.25,12.75) | 21 | 264 | 5.8(5.13,6.55) |
| Complications of pregnancy, childbirth, puerperium | 22 | 377 | 85.18(76.79,94.22) | 25 | 26 | 40.43(14.84,87.99) |
| Tuberculosis | 23 | 325 | 6.07(5.43,6.77) | 22 | 9 | 4.64(4.02,5.34) |
| Certain conditions originating in perinatal period | 24 | 29 | 12.58(8.42,18.06) | 23 | 6 | 2.83(1.85,4.14) |
| Syphilis | 25 | 13 | 2.44(1.3,4.17) | 24 | 196 | 1.7(0.78,3.22) |

Abbreviations: SMR, standardized mortality ratios; CI, confidence interval.
<sup>1</sup>Causes of death are ranked by the number of observed deaths.
<sup>2</sup>The SMR is calculated as the ratio of deaths observed in cancer patients to the deaths expected in the general population, adjusted for age, sex, race, and year. An SMR above 1 means a higher risk of death for the cancer patients.

SMRs for the four major non-cancer causes of death were stratified by age at diagnosis and time since diagnosis (Figure 4). Younger patients generally had the highest SMRs. Cancer patients had a significantly elevated risk of death from chronic liver disease and cirrhosis compared to the general population, with the highest SMRs observed among hepatocellular carcinoma patients. Chronic liver disease and cirrhosis were also associated with higher SMRs among patients diagnosed with ovarian, gastric, pancreatic, and lung cancers. Hematologic malignancies (e.g., non-Hodgkin lymphoma, leukemia, multiple myeloma) were linked to higher SMRs for deaths from infectious and parasitic diseases, including HIV. Lung and liver cancer patients also had elevated SMRs for infectious and parasitic diseases. Among younger patients, those with ovarian, breast, gastric, and pancreatic cancers had the highest SMRs for suicide and self-inflicted injuries (Figure 4A). Patients with brain and neurologic malignancies were most likely to die from non-cancer causes within the first year after diagnosis compared to patients with other cancers. Among patients who died from infectious diseases and HIV, those with pharyngeal or hematologic malignancies had the highest SMRs. Patients with pancreatic cancer had the highest SMR for suicide and self-inflicted injuries in the first year after diagnosis. In addition, cervical cancer patients showed a continued increase in the risk of suicide and self-injury over time, with the highest SMR observed more than 10 years after diagnosis (Figure 4B).

**Figure 4.**
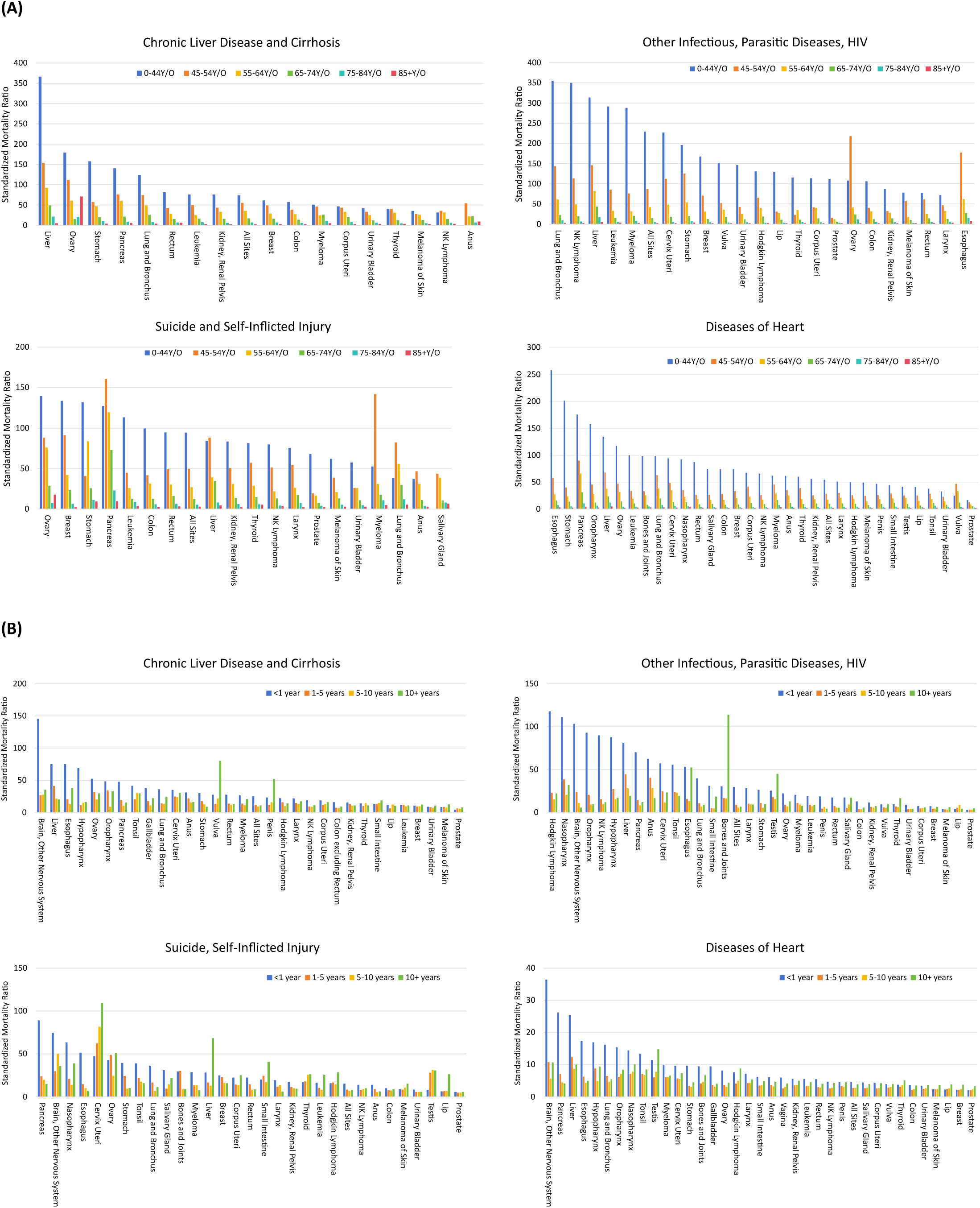
Standardized mortality ratios (SMRs) for the leading causes of non-cancer death among cancer patients diagnosed between 2000 and 2021 by (A) age group and (B) time periods after diagnosis.

### Emerging Trends in Cause of Death: A Comparative Analysis of 1975-1999 and 2000-2021

For comparison, we also analyzed cause-of-death data for cancer patients registered with SEER between 1975 and 1999. Detailed analytical methods and results are provided in the supplementary material. In contrast to the trends observed during 1975–1999, our study of cancer patients diagnosed between 2000 and 2021 revealed several emerging trends in the causes of cancer-related deaths in the 21st century.

Overall, although the trends in the proportions of different causes of death were largely consistent across the two periods, the slope was steeper in the 21st century. From 2000 to 2021, the slope for index cancer death cause was –1.58, compared to –1.09 between 1975 and 1999. Similarly, the slopes for non-cancer death causes were 1.36 and 0.86 for 2000-2021 and 1975-1999, respectively (Figure 1A, Supplemental Figure S1A). Specifically, between 2000 and 2021, index cancer mortality rates for cancers such as brain malignancies, pancreatic cancer, and gallbladder cancer remained consistently high, similar to historical trends, while mortality rates for lung, liver, esophageal cancers, and multiple myeloma declined. Notably, the mortality rate for leukemia showed an even more pronounced decline. While non-index cancer deaths were not the primary cause of death overall, a high proportion of non-index cancer deaths was observed in patients with head and neck cancers (e.g., oral cavity, oropharynx), vaginal cancer, and small intestine cancer. Particularly in oropharyngeal cancer, the proportion of non-index cancer deaths ranged from 50% to 60%, a pattern consistent across both the 1975-1999 and 2000-2021 periods. Nevertheless, the proportion of non-index cancer deaths from small intestine cancer increased significantly during the 2000-2021 period. Impressively, the characteristics of non-cancer causes of death among cancer patients differed markedly between the two periods (Figure 2, Supplemental Figure S2).

A notable trend in the 21st century has been the increased risk of various non-cancer causes of death, with most causes showing higher SMR compared to the previous century. From 1975 to 1999, heart disease was the leading non-cancer cause of death, with an SMR of 1.84. By 2000-2021, the SMR for heart disease had risen to 3.11. One striking change was the significant increase in risk for Alzheimer’s disease, with its SMR rising from 0.41 in 1975-1999 to 1.84 in 2000-2021. Moreover, from 1975 to 1999, cancer patients did not exhibit a statistically significant difference in the risk of death from hypertension (without heart disease) or from symptoms, signs, and ill-defined conditions compared to the general population. However, from 2000 to 2021, the risks for both conditions among cancer patients surpassed those of the general population, with SMR greater than 1. Notably, the risk of death from other infectious and parasitic diseases, including HIV, decreased over time, with the SMR dropping from 16.25 in 1975-1999 to 9.98 in 2000-2021 (Table 2). After stratifying cancer patients who died from non-cancer causes by age and follow-up time after diagnosis, it was found that, overall, the proportion of younger patients dying from non-bacterial infections has significantly decreased in the 21st century, particularly among those aged 25 to 49. In contrast, the proportion of deaths due to accidents and adverse effects has correspondingly increased. Among older patients, the proportion of deaths from heart disease has slightly decreased (Figure 3A, Supplemental Figure S3A). The trends in the proportion of non-cancer causes of death over time after diagnosis are generally consistent between the two periods, with a slight decline in deaths from heart disease and a slight increase in deaths from other causes (Figure 3B, Supplemental Figure S3B). Notably, the SMR charts for the four leading non-cancer causes of death reveal that in the 21st century, the mortality risks for chronic liver disease and cirrhosis, suicide and self-inflicted injury, and diseases of heart generally increased across all age groups and follow-up periods. However, a different trend was observed for other infectious and parasitic diseases, including HIV, where the mortality risk exhibited a broad decline across follow-up periods compared to the years 1975-1999 (Figure 4, Supplemental Figure S4).

## Discussion

We analyzed the causes of death for 3,364,903 cancer patients registered with SEER between 2000 and 2021, identifying the characteristics of the cause-of-death spectrum among cancer patients in the first two decades of the 21st century. The causes of death varied by cancer type, time since diagnosis, and age at diagnosis.

Certain types of cancer, such as malignant brain tumors, pancreatic cancer, and gallbladder cancer, persistently show high mortality rates directly linked to the primary disease. This highlights the urgent need for more effective prevention strategies, early detection through screening and surveillance, and the development of new treatment options. Zaorsky et al. have reported that cancers including liver, pancreatic, lung, nasopharyngeal, esophageal, malignant brain tumors, and multiple myeloma remained among those with the highest rates of death due to index cancer for patients diagnosed between 1973 and 2012^9^. Our study corroborates this finding, revealing a significant proportion of deaths attributable to index cancers within these types. However, we observed a notable decline in the proportion of deaths from index cancers for lung, liver, nasopharyngeal, esophageal cancers, and multiple myeloma in the 21st century. This trend indicates a substantial shift in the cancer mortality profile, likely attributable to advancements in cancer management strategies in recent years. The progress in cancer management strategies in the new century is heavily influenced by personalized medicine. Although the concept of personalized medicine emerged in the late 20th century^18^, it has rapidly advanced and matured in the early 21st century due to developments in genomics, molecular biology, and informatics^19,20^. Precision oncology, a critical aspect of personalized medicine, has become a central focus in contemporary cancer treatment^21^. Significant strides have been made in the clinical application of targeted therapies and immunotherapies. For instance, targeted agents have markedly improved survival rates in lung cancer patients^22,23^, while novel immunomodulatory drugs, such as proteasome inhibitors, have notably enhanced remission rates and overall survival in multiple myeloma patients^24^. In esophageal cancer, trastuzumab has significantly improved outcomes in HER2-positive patients^25^. The decline in deaths from liver cancer-index cancers may be attributed to new staging and scoring systems, such as the Barcelona Clinic Liver Cancer (BCLC) staging system^26–28^, the Milan criteria^29^, and the Model for End-Stage Liver Disease (MELD) score^30^, which are crucial for evaluating liver cancer patients for transplantation. Advances in locoregional therapies, including radiofrequency ablation (RFA) and transarterial chemoembolization (TACE), have also provided new treatment options for patients with unresectable liver cancer^31^. Additionally, targeted drugs like Sorafenib have significantly extended survival in advanced liver cancer cases^32^. The reduction in deaths from nasopharyngeal carcinoma may be linked to improvements in radiation therapy techniques^33^. Despite these advances, the proportion of deaths from index cancers remains high, suggesting that current and emerging treatment and surveillance strategies may only benefit a subset of the patient population.

Our study found that the proportion of non-index cancer deaths exceeded the proportion of index cancer deaths for oral cavity, oropharyngeal, vaginal, and small intestine cancers. Additionally, the absolute number of non-index cancer deaths was highest among patients with prostate, breast, and lung cancers. This observation highlights the significance of non-index cancers among cancer survivors. Similarly, Makoto et al. reported a high proportion of non-index cancer deaths for oral cavity and pharyngeal cancers in a Japanese cancer cohort from 1985 to 2014^34^. This phenomenon may be attributable to common carcinogenic factors, such as genetic susceptibility, environmental influences, or lifestyle choices, as well as treatment-related factors. For instance, oral cavity and oropharyngeal cancers are linked to prolonged exposure to carcinogens like tobacco, alcohol^35^, and HPV infection^36^, which can lead to simultaneous or sequential cancers at multiple sites and thereby increase the risk of non-index cancers. While chemotherapy, radiation therapy, and other cancer treatments are effective for controlling the primary cancer, they may inadvertently induce secondary cancers. For example, radiotherapy can elevate the risk of cancers at sites outside the treated area, and chemotherapy may heighten the risk of cancers in other organs due to its systemic effects^37,38^. Therefore, patients with these cancers would benefit from vigilant surveillance and early screening for non-index cancers. Comprehensive physical examinations and targeted imaging for all cancer survivors are essential for the early detection of potential non-index cancers. Endoscopic screening following a diagnosis of oral cancer has been shown to significantly enhance the early detection of second primary esophageal cancer and reduce all-cause mortality from oral cancer^39^. Additionally, optimizing cancer treatment regimens is crucial. Treatment plans should aim to minimize damage to healthy tissues by adjusting radiotherapy doses and employing more selective chemotherapeutic agents. Lifestyle modifications, including smoking cessation, reducing alcohol intake, and minimizing exposure to carcinogens, can also help mitigate the risk of non-index cancers. Future studies should investigate the non-index cancers most likely to occur and their associated risk factors in patients with the aforementioned cancer types. By identifying these specific non-index cancers and their risk factors, interventions can be more precisely tailored to reduce mortality from non-index cancers.

This study analyzed deaths from non-cancer causes in cancer patients between 2000 and 2021, revealing the significant impact of non-cancer causes of death such as heart disease, chronic liver disease and cirrhosis in specific groups of cancer patients. Notably, the SMR for non-cancer causes of death was significantly higher in younger patients compared to other age groups, indicating that younger cancer patients may face an elevated risk of non-cancer deaths following treatment. Our results corroborate previous findings regarding the predominance of heart disease in cancer patients^9^. Cancer patients have a higher risk of death from heart disease compared to the general population, with this risk being particularly pronounced in patients younger than 45 years. This heightened risk may be related to shared risk factors between cardiovascular disease and cancer^40^, the cancer itself, and the complex effects of cancer treatment on the cardiovascular system. Certain cancer treatments, notably anthracyclines, immunotherapies, and targeted therapies, have been associated with increased risks of myocardial injury, arrhythmias, and heart failure^41^. Inflammation and dysregulation of the immune system, common in cancer patients, can also accelerate atherosclerosis, further increasing the risk of cardiac disease^42^. The interactions between heart disease and cancer are complex^43^. Additionally, epidemiologic studies have demonstrated an increased risk of cancer in patients with heart failure^44^. The risk of heart disease extends beyond the treatment period; with prolonged survival after diagnosis, patients continue to face long-term cardiovascular risks. Our study confirms that the risk of death from heart disease increases with time after diagnosis, as evidenced by the work of Kelsey C et al^45^. Furthermore, our study examined the risk of death from chronic liver disease and cirrhosis in cancer patients. Previous epidemiologic research has highlighted a significant increase in SMR for chronic liver disease and cirrhosis in patients with digestive system cancers^46^. Liver cancer patients are at an elevated risk of developing chronic liver disease and cirrhosis due to direct liver damage. However, this increased risk is also notably significant in patients with other cancers, including ovarian, gastric, pancreatic, and lung cancers. This phenomenon can be attributed to various mechanisms. Cancer treatments may induce liver toxicity, leading to chronic damage and fibrosis that can progress to chronic liver disease and cirrhosis. For example, certain commonly used targeted drugs^47^ and immunotherapeutic agents^48^ have been shown to have high hepatotoxicity, contributing to the development of chronic liver disease. Additionally, the systemic inflammatory response and metabolic changes^49^ associated with cancer may exacerbate liver injury, increasing the risk of death from chronic liver disease. These findings underscore the importance of monitoring and protecting liver function during cancer treatment, especially for patients with cancers other than hepatocellular carcinoma. The SMR for other infectious diseases, parasitic diseases, and HIV is significantly elevated in certain cancer patients, particularly those with hematopoietic malignancies. This elevation may be related to impaired immune function, which makes these patients more susceptible to infections. Studies have shown that immunosuppressive agents used in cellular therapy^50^, immunotherapy, and molecularly targeted drugs^51^ can increase susceptibility to infectious diseases. Consequently, these patients are not only more prone to infections but also more likely to experience severe complications, which heightens their risk of death. Higher SMRs for infections have also been observed in patients with lung and liver cancers, suggesting these cancer types may be associated with a greater risk of infection. Enhanced infection prevention and early treatment measures are crucial for these patient groups to reduce mortality risk. Lastly, patients with pancreatic cancer exhibited the highest SMR for suicide and self-injury within the first year after diagnosis, likely related to the disease’s high lethality and associated psychological stress^52^. Conversely, patients with cervical cancer showed a significantly increased risk of suicide and self-injury more than 10 years after diagnosis, highlighting the need for ongoing mental health support for long-term survivors. This suggests that the quality of life and mental health issues of long-term cervical cancer survivors should not be overlooked^53^. These findings emphasize the importance of psychological support and mental health interventions during cancer treatment and follow-up. Additionally, the increased risk of death from Alzheimer’s disease and hypertension in cancer patients in the 21st century may be associated with improved cancer treatments and an aging population, suggesting the need for comprehensive health management strategies to address the multiple health risks of cancer patients.

Our study findings have revealed significant characteristics in the causes of death among cancer patients between 2000 and 2021, in contrast to the trends observed from 1975 to 1999. The declining trends in index cancer mortality rates for specific cancer types, such as lung and liver cancer, in the 21st century may reflect advancements in cancer treatment and management, as well as the broader implementation of early diagnostic techniques, leading to improved survival rates. The widespread increase in the risk of various non-cancer causes of death underscores the importance of prioritizing patients’ overall health rather than focusing exclusively on the cancer itself. The persistently higher rates of non-index cancer mortality in certain cancers, such as head and neck cancers and small intestine cancer, across both periods suggest that these patients may profit significantly from second cancer screenings.

Despite the remarkable advancements in cancer science research during this century, cancer continues to pose a substantial threat to global health, suggesting a gap between foundational scientific research and clinical translation. Consequently, a thorough analysis of causes of death is crucial for evaluating the survival benefits of cancer patients from a macro perspective. This study offers a comprehensive, population-based analysis of causes of death among cancer patients, reflecting the most current data available. Admittedly, it is important to acknowledge certain limitations. First, inherent biases, such as selection bias and information bias, may arise due to the retrospective nature of the analysis. Second, the classification of causes of death may present challenges, particularly when complete or detailed medical records are lacking; for instance, a patient’s cause of death might be inaccurately categorized as “other causes” or deemed undetermined. In addition, a longer follow-up period allows for the accumulation of additional non-index cancer deaths and non-cancer deaths, which may lead to a gradual increase in the proportion of such deaths over calendar years. Finally, this study relies on data from the SEER database, which is representative of the U.S. population, thus presenting limitations regarding external generalizability. However, as economic and healthcare standards improve in developing countries, those with higher levels of development can leverage the findings of this study to guide the formulation of medium– and long-term cancer control and public health policies. To further enhance the generalizability of future research, we recommend incorporating data from a broader range of races and regions. Furthermore, many countries lack systematic records of causes of death, which are crucial for clinical translational research and public health policy development. Therefore, establishing comprehensive databases on cancer-related deaths is both urgent and essential for global health development.

## Conclusion

This study highlights the trends in three categories of death causes—index cancer causes, non-index cancer causes, and non-cancer causes—among cancer patients during the first two decades of the 21st century, along with the specific cause-of-death characteristics for each cancer site. The persistently high mortality rates from index cancers of the brain, pancreas, and gallbladder underscore the urgent need for more effective prevention, screening, and treatment strategies. Notably, while mortality rates from index cancers of the lung, liver, nasopharynx, esophagus, and multiple myeloma were historically high, they have shown a decline in the first two decades of the 21st century, reflecting advancements in cancer management. Conversely, mortality rates from non-index cancers in patients with oral cavity, oropharyngeal, vaginal, and small intestine cancers exceeded those from index cancers, indicating that these patients might benefit from enhanced surveillance and early screening for non-index cancers. Additionally, several non-cancer causes of death, including heart disease, chronic liver disease and cirrhosis, remain significant contributors to mortality among cancer patients. The trends revealed in this study provide crucial insights for the future management of cancer patients. As mortality rates for certain index cancers continue to decline, the widespread implementation of cancer screening and early diagnosis should be further emphasized to reduce mortality across more cancer types. Meanwhile, for cancers with high mortality from non-index cancers, increased surveillance and preventive strategies for non-index cancers are essential to help patients avoid potential secondary cancer threats. Furthermore, the prominence of non-cancer causes of death, such as heart disease and chronic liver disease, underscores the necessity of a holistic approach to health management in cancer patients. Future treatment strategies should be more comprehensive to improve the overall quality of life for cancer patients.

## Data Availability

All data produced in the present work are contained in the manuscript

https://seer.cancer.gov/data-software/documentation/seerstat/nov2021/

## ACKNOWLEDGMENTS

This work was partially supported by the National Natural Science Foundation of China (81602167; 82372617 and 81972658; 81803636), the Hunan Provincial Natural Science Foundation of China (2017JJ3494 and 2021JJ31100), the Open Project of Xiangjiang Laboratory (23XJ03001), the Science and Technology Program Foundation of Changsha City (kq2004085), the Fundamental Research Funds for the Hunan Provincial Innovation Foundation for Postgraduate (CX20230328), the Guangdong Basic and Applied Basic Research Foundation (2024B1515020090 and 2023A1515012683; 2024A1515030038; 2023B1212060013 and 2020B1212030004), and the Basic and Applied Basic Research of Guangzhou Municipal Basic Research Plan (2024A03J0845 and 2023A04J2098). In addition, we appreciate laboratory members for thoughtful suggestions and comments on the manuscript.

