## Supplemental Figure for "Causes of Death among Cancer Patients: Emerging Trends in the 21st Century"

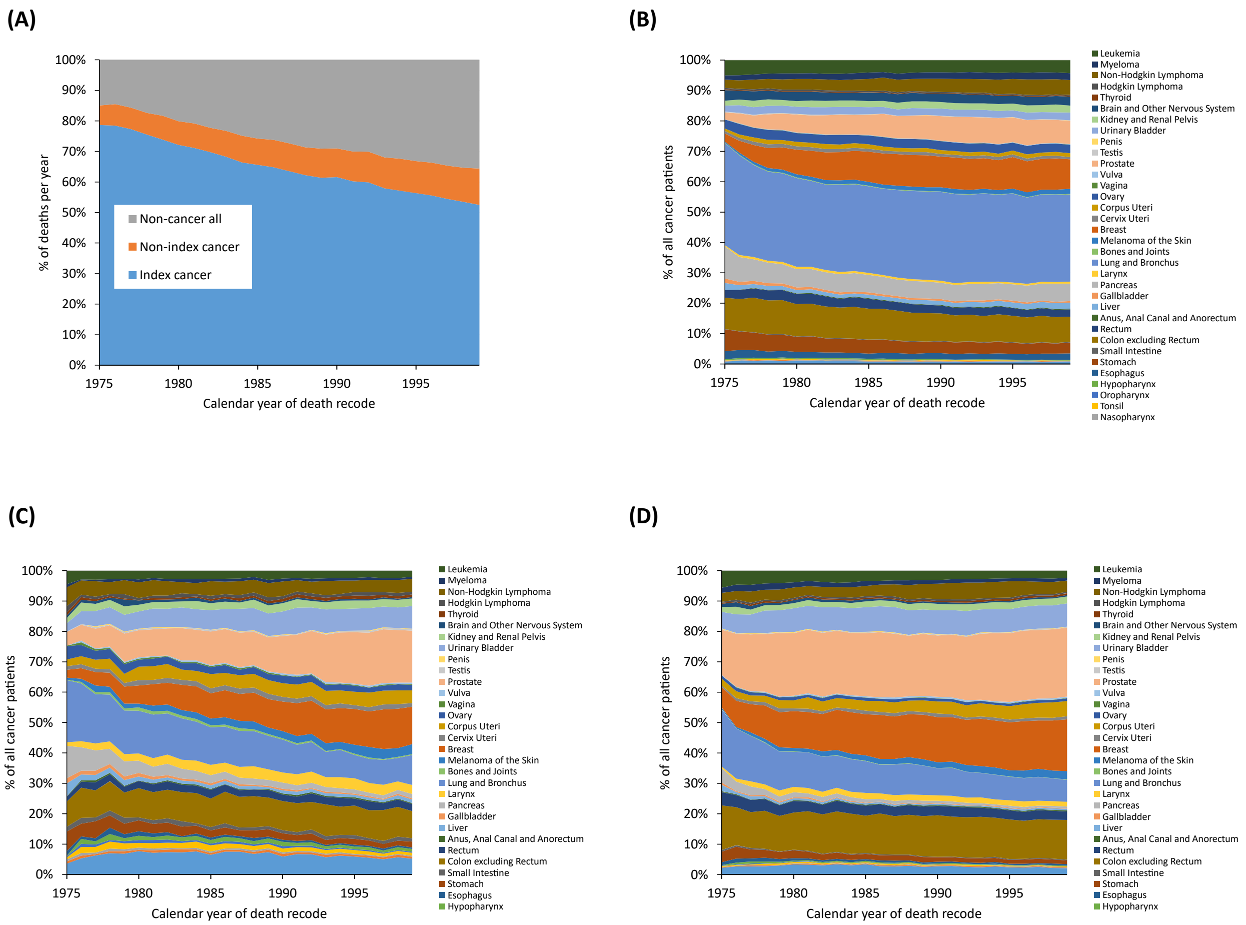


**Supplemental Figure S1.** Overall trends in causes of death among cancer patients diagnosed between 1975 and 1999 . (A) Proportional graph of cancer patients' deaths due to index cancers (black), non-index cancers (yellow), and non-cancer causes (green) from 1975 to 1999. The proportion of index-cancer deaths has been continuously decreasing, while deaths due to non-cancer causes have increased. Proportional graph of deaths caused by (B) index cancers, (C) non-index cancers, and (D) non-cancer causes, stratified by various types of cancer.

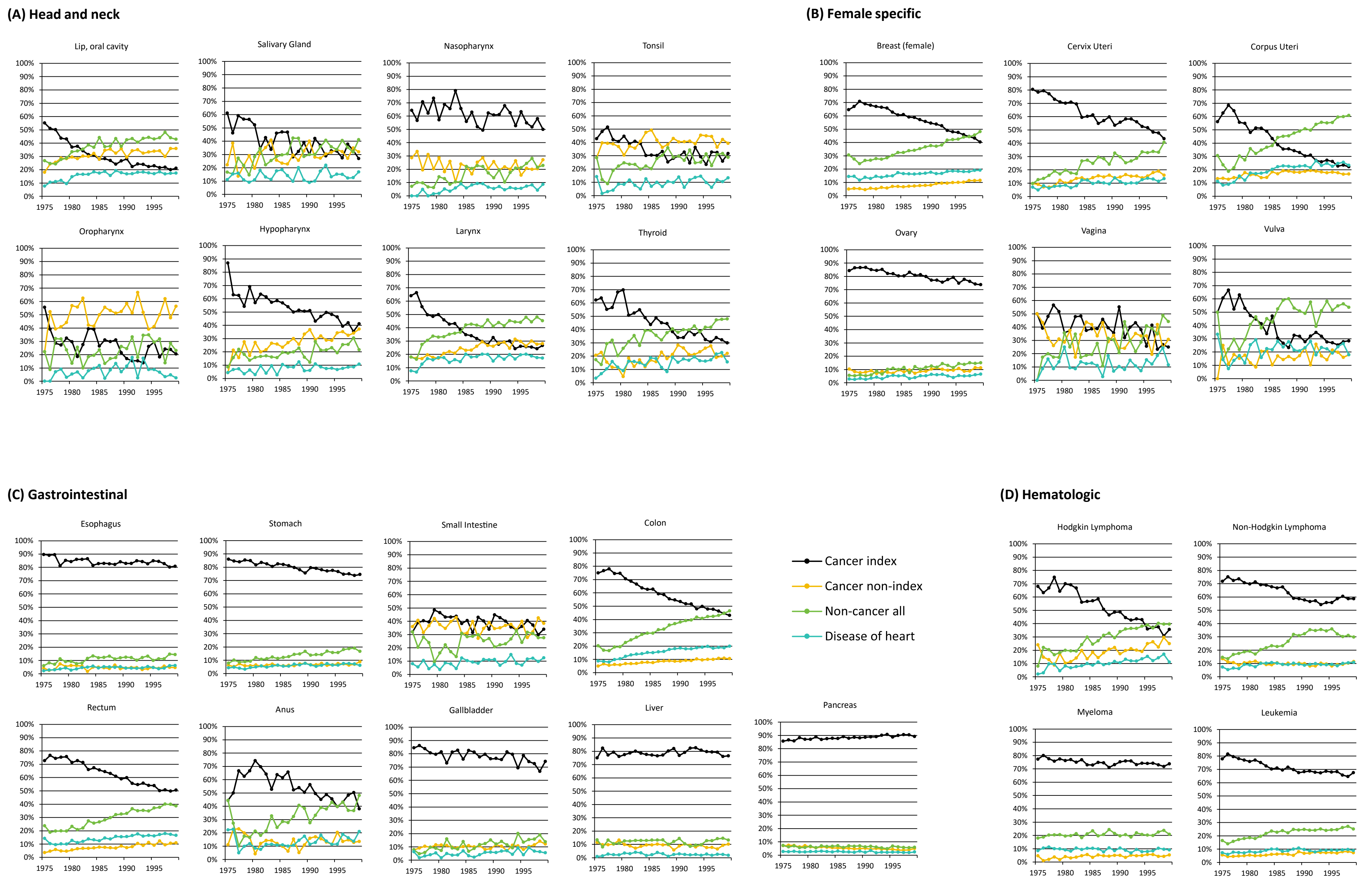
(A) Head and neck
Lip, oral cavity
Salivary Gland
Nasopharynx
Tonsil
Oropharynx
Hypopharynx
Larynx
Thyroid
(B) Female specific
Breast (female)
Cervix Uteri
Corpus Uteri
Ovary
Vagina
Vulva
(C) Gastrointestinal
Esophagus
Stomach
Small Intestine
Colon
Rectum
Anus
Gallbladder
Liver
Pancreas
(D) Hematologic
Hodgkin Lymphoma
Non-Hodgkin Lymphoma
Myeloma
Leukemia
Cancer index
Cancer non-index
Non-cancer all
Disease of heart

**(E) Male specific**

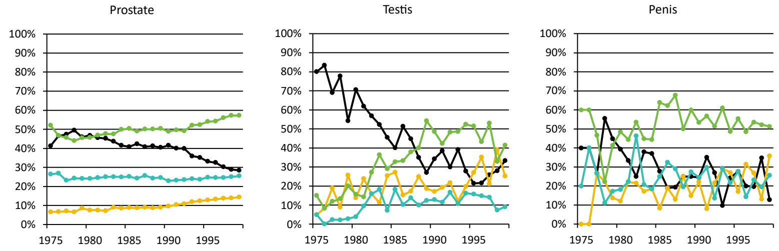


**(F) Other**

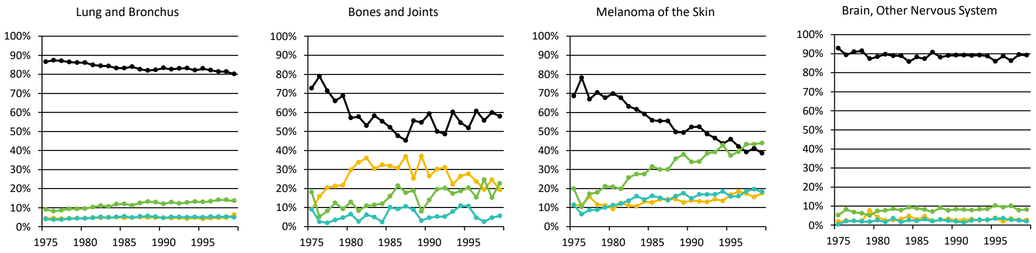


**(G) Genitourinary**

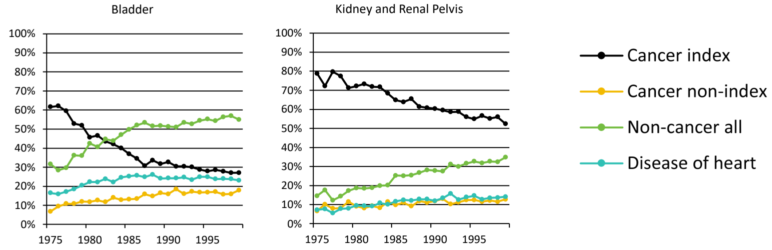


**Supplemental Figure S2.** Proportion of cause of death versus year of death among cancer patients diagnosed between 1975 and 1999 for various cancers. Death was characterized as due to “index-cancer,” (black lines), “non-index cancer” (yellow lines), and “noncancer death” (green lines). Year of death refers to the year in which the death occurred.

(A)

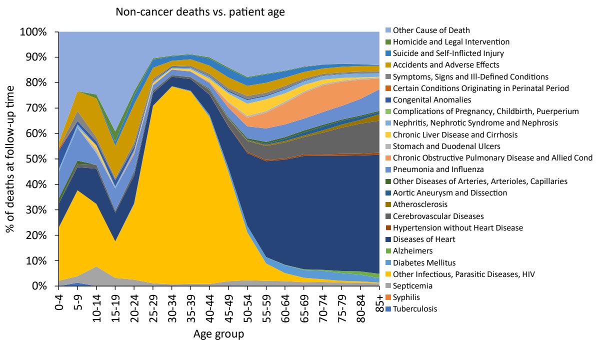


(B)

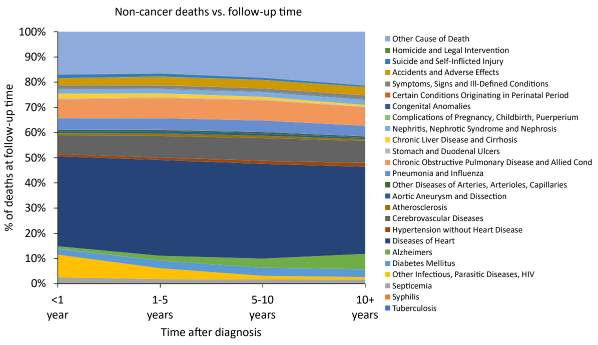


**Supplemental Figure S3.** Overall trends of the proportion of non-cancer causes of death among cancer patients. Proportional graphs of death due to non-cancer causes among cancer patients diagnosed between 1975 and 1999 by (A) patient age and (B) time after diagnosis, stratified by various non-cancer causes.

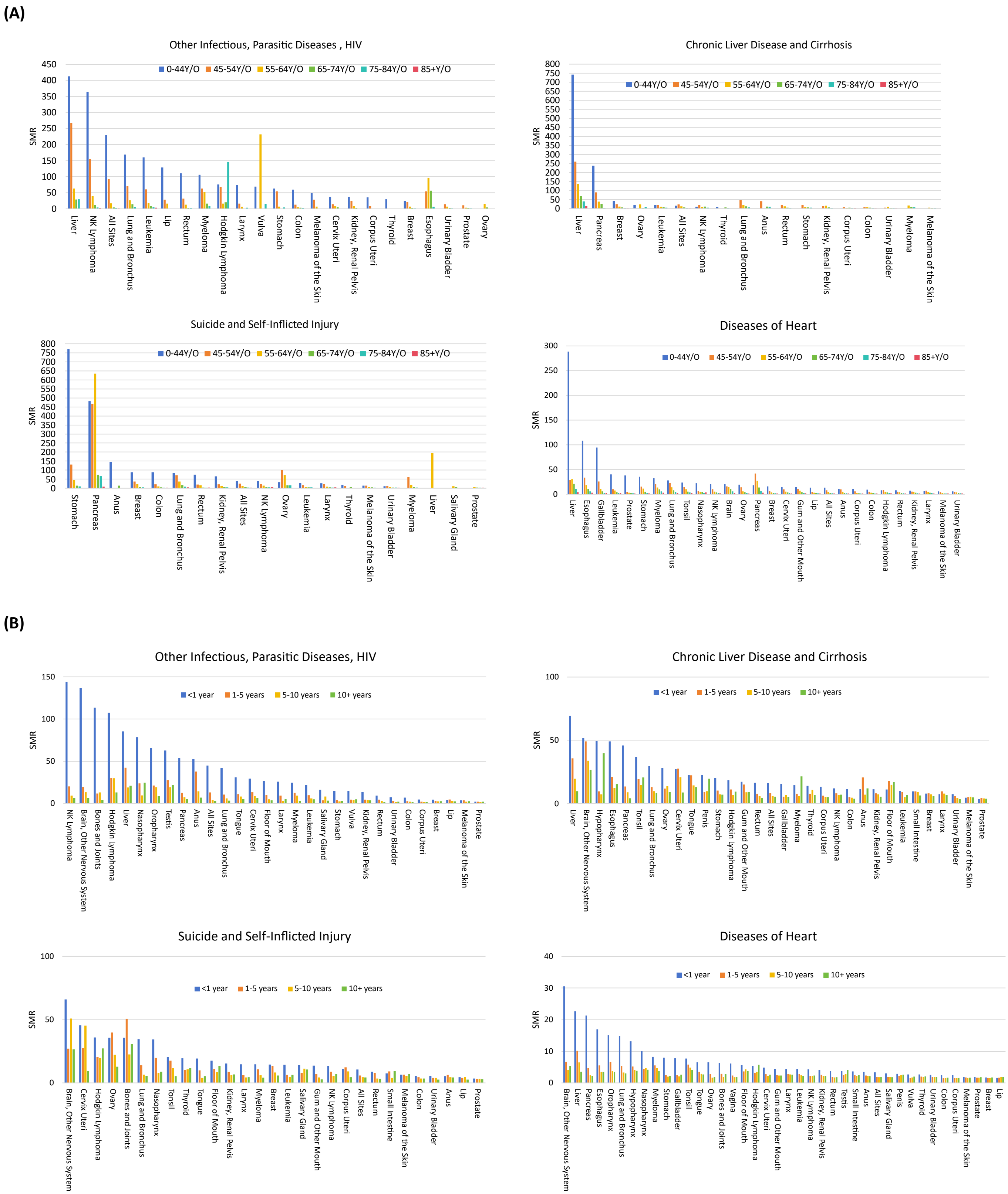


**Supplemental Figure S4.** Standardized mortality ratios (SMRs) for the leading causes of non-cancer death among cancer patients diagnosed between 1975 and 1999 by (A) age group and (B) time periods after diagnosis.
