## Supplemental Methods for "Causes of Death among Cancer Patients: Emerging Trends in the 21st Century"

**Supplementary Methods for the Analysis of Causes of Death Among Cancer Patients from 1975 to 1999:**

We use SEER*Stat software (version 8.4.3) to extract data on cancer patients diagnosed with cancers from January 1, 1975, to December 31, 1999 from the SEER 8 registry (2023 submission), which includes approximately 8.3% of the U.S. population^1-2^. The definition of variables and the methods of data analysis are consistent with those in the manuscript.

1. Surveillance Research Program, National Cancer Institute SEER*Stat software (seer.cancer.gov/seerstat) version 8.3.4
2. Surveillance, Epidemiology, and End Results (SEER) Program (www.seer.cancer.gov) SEER*Stat Database: Incidence - SEER Research Data, 8 Registries, Nov 2023 Sub (1975-2021) - Linked To County Attributes - Time Dependent (1990-2022) Income/Rurality, 1969-2022 Counties, National Cancer Institute, DCCPS, Surveillance Research Program, released April 2024, based on the November 2023 submission.
