## Supplemental Table 1 for "Causes of Death among Cancer Patients: Emerging Trends in the 21st Century"

**Supplemental Table S1.** The non-cancer causes of death using SEER’s standard 1969+ Recode

| **Non-Cancer Causes of Death*** | **ICD-10*** |
| --- | --- |
| Tuberculosis | A15-A19 |
| Syphilis | A50-A53 |
| Human Immunodeficiency Virus (HIV) (1987+)* | B20-B24 |
| Septicemia | A40-A41 |
| Other Infectious and Parasitic Diseases* | A00-A08, A20-A33, A35-A39, A42-A49, A54-B19, B25-B99 |
| Diabetes Mellitus | E10-E14 |
| Alzheimers (ICD-9 and 10 only) | G30 |
| Diseases of Heart | I00-I09, I11, I13, I20-I51 |
| Hypertension without Heart Disease | I10, I12 |
| Cerebrovascular Diseases | I60-I69 |
| Atherosclerosis | I70 |
| Aortic Aneurysm and Dissection | I71 |
| Other Diseases of Arteries, Arterioles, Capillaries | I72-I78 |
| Pneumonia and Influenza | J09-J18 |
| Chronic Obstructive Pulmonary Disease and Allied Cond | J40-J47 |
| Stomach and Duodenal Ulcers | K25-K28 |
| Chronic Liver Disease and Cirrhosis | K70, K73-K74 |
| Nephritis, Nephrotic Syndrome and Nephrosis | N00-N07, N17-N19, N25-N27 |
| Complications of Pregnancy, Childbirth, Puerperium | A34, O00-O95, O98-O99 |
| Congenital Anomalies | Q00-Q99 |
| Certain Conditions Originating in Perinatal Period | P00-P96 |
| Symptoms, Signs and Ill-Defined Conditions | R00-R99 |
| Accidents and Adverse Effects | V01-X59, Y85-Y86 |
| Suicide and Self-Inflicted Injury | U03, X60-X84, Y87.0 |
| Homicide and Legal Intervention | U01-U02, X85-Y09, Y35, Y87.1, Y89.0 |
| COVID-19 (2020+) [6](https://seer.cancer.gov/codrecode/1969_d03012018/index.html" \l "footnotes) | U07.1 |
| Other Cause of Death |  |

*Non-Cancer Causes of Death and ICD-0-10 columns adapted from SEER: [SEER Cause of Death Recode 1969+ (03/01/2018) (cancer.gov)](https://seer.cancer.gov/codrecode/1969_d03012018/index.html)
