## Supplemental Table 2 for "Causes of Death among Cancer Patients: Emerging Trends in the 21st Century"

**Supplemental Table S2**: Definition of index-cancer

The coding of cause of death due to the index cancer may be imperfect for certain diseases. In order to address these inconsistencies, deaths attributed to specific subsites of locoregional cancer were included as part of the index cancer.

| **Locoregional cancer** | **Specific subsites** |
| --- | --- |
| Nasopharynx | Other oral cavity and pharynx cancer |
| Tonsil | Oropharynx  Other oral cavity and pharynx cancer |
| Oropharynx | Other oral cavity and pharynx cancer |
| Hypopharynx | Other oral cavity and pharynx cancer  Larynx |
| Esophageal | Stomach |
| Stomach | Esophageal |
| Colon | rectum and rectosigmoid junction  Anus, anal canal, and anorectum |
| Rectum | Colon excluding rectum  Anus, anal canal, and anorectum |
| Anal | Colon excluding rectum  Rectum and rectosigmoid junction |
| Liver | Intrahepatic bile duct  Gallbladder  Other biliary sites |
| Gallbladder | Liver  Intrahepatic bile duct  Other biliary sites |
| Pancreas | Other biliary  Liver |
| Larynx | Tonsil  Oropharynx  Hypopharynx  Other oral cavity and pharynx cancer |
| Lung and bronchus | Pleura  Trachea, mediastinum and other respiratory organs |
| Cervix uteri | Corpus uteri  Uterus, Nos |
| Corpus uteri | Cervix uteri  Uterus, Nos |
| Vulva | Vagina |
| Urinary bladder | Renal pelvis  Ureter |
